# Perioperative risk of non-cardiac surgery in patients with asymptomatic significant aortic stenosis: a 10-year retrospective study

**DOI:** 10.1101/2023.09.14.23295589

**Authors:** Minjung Bak, Seung-Hwa Lee, Sung-Ji Park, Jungchan Park, Jihoon Kim, Darae Kim, Eun Kyoung Kim, Sung-A Chang, Sang-Chol Lee, Seung Woo Park

**Author notes:** Corresponding Author: Sung-Ji Park, MD, PhD Division of Cardiology, Department of Internal Medicine, Cardiovascular Imaging Center, Heart Vascular Stroke Institute, Samsung Medical Center, Sungkyunkwan University School of Medicine, 81 Irwon-ro, Gangnam-gu, Seoul, Republic of Korea. Minjung Bak and Seung-Hwa Lee contributed equally to this work.

## Abstract

**BACKGROUND:** Aortic valve stenosis (AS) is a representative geriatric disease, and there is an anticipated rise in the number of patients requiring non-cardiac surgeries in patients with AS. However, there is still a lack of research on the primary predictors of non-cardiac perioperative complications in patients with asymptomatic significant AS.

**METHOD AND RESULT:** Among the cohort of non-cardiac surgeries under general anesthesia, with an intermediate-to-high risk of surgery from 2011 to 2019, at Samsung Medical Center, 221 patients were identified to have asymptomatic significant AS. First, to examine the impact of significant AS on perioperative adverse events, the occurrences of Major Adverse Cardiovascular Events (MACE) and Perioperative Adverse Cardiovascular Events (PACE) were compared between patients with asymptomatic significant AS and the control group. Second, to identify the factors influencing the perioperative adverse events in patients with asymptomatic significant AS, multivariable logistic regression model was used.

There was no significant difference between the three groups consisting of control group, moderate asymptomatic AS group, and severe asymptomatic AS group in the event rate of MACE (p value 0.971) and PACE (p value 0.185). Cardiac damage stage was significant risk factor of MACE (OR 9.241, 95% CI: 1.314 - 64.976, p-value = 0.025) and PACE (OR 2.199, 95% CI: 1.055 - 4.584, p-value = 0.036).

**CONCLUSION:** There was no significant difference in major post-operative cardiovascular events between patients with asymptomatic significant AS and the control group. Advanced cardiac damage stage in significant AS is a key factor in perioperative risk of non-cardiac surgery.

**CLINICAL PERSPECTIVE:** *What Is New?:* Advanced cardiac damage is a major risk factor for perioperative complications in non-cardiac surgeries of patients with asymptomatic significant AS.

*What Are the Clinical Implications?:* Patients with asymptomatic significant AS showed no significant difference in perioperative complications compared to the control group, but caution is needed in those with advanced cardiac damage, as they are more likely to experience adverse events.

## INTRODUCTION

The increasing life expectancy has resulted in a growing elderly population, consequently leading to a rise in the number of patients with comorbidities requiring non-cardiac surgeries. Within this population, aortic valve stenosis (AS) is a representative geriatric disease, and the demand for surgical interventions in patients with significant AS is anticipated to escalate.^1,2^ In patients with AS, increased afterload results in concentric hypertrophy, which leads to diastolic dysfunction and decreased coronary flow reserve.^3^ In addition, patients with AS have many underlying diseases, such as coronary artery disease (CAD), hypertension, and diabetes. The pathophysiology of AS, combined with these comorbidities, renders patients with AS particularly vulnerable to hemodynamic changes that occur during anesthesia. Despite advancements in anesthesia technology that have contributed to a decline in perioperative cardiovascular events, significant AS continues to be recognized as a significant risk factor for non-cardiac surgeries.^4–6^

Nevertheless, there is a need for updated studies focusing on the perioperative risk of significant AS with surgeries performed after 2010, as previous research has mainly concentrated on surgeries conducted before that time.^4,5,7^ Furthermore, despite recognition that operative risk is elevated in individuals with AS and coexisting congestive heart failure, the relationship between the recently proposed cardiac damage stage and operative risk in AS patients has not yet been established.^5,8–10^ Additionally, although previous studies have indicated a high operative risk for patients with typical AS symptoms,^5,7,10,11^ only a limited number have thoroughly examined the differences between asymptomatic AS patients and controls without AS.^12^ Hence, the objective of this study was to investigate the key risk factors associated with non-cardiac surgery in asymptomatic patients with significant AS and compare the surgical outcomes between this group and a control group in the present era.

## METHODS

### Patient Selection

From 2011 to 2019, 203,788 patients underwent non-cardiac surgery at the Samsung Medical Center (SMC). The above patients were registered in the SMC-non-cardiac operation registry (KCT 0006363). After excluding patients with a history of aortic valve replacement, patients without general anesthesia, and low-risk patients, 113,078 patients with intermediate to high-risk non-cardiac surgery were included in the analysis. The surgical risk was stratified according to the American College of Cardiology/American Heart Association Guidelines and the European Society of Cardiology/European Society of Anesthesiology guidelines on non-cardiac surgery on Perioperative Cardiovascular Evaluation and Management of Patients Undergoing Non-cardiac Surgery.^13,14^ Significant AS was defined as patients with more than a moderate degree of AS who were diagnosed before surgery, or that was confirmed on preoperative echocardiography according to the current guidelines.^15^ Patients with moderate AS and patients with severe AS were analyzed together in this study because moderate AS has been shown to have a poor prognosis similar to severe AS and accounts for a significant proportion of patients undergoing surgery.^16^

There were 223 patients with significant AS among this population according to the criteria. Among these patients, 178 patients had moderate AS, and 45 patients had severe AS. In a medical chart review, among these 223 patients, 2 patients with severe AS had symptoms. These two patients complained of dyspnea of New York Heart Association class II and III, respectively, and there was no explanation for the symptoms other than severe AS. These two patients were excluded from the analysis. To compare the prognosis between patients with asymptomatic significant AS and those without significant AS, 218 asymptomatic significant AS patients and 436 control groups were selected through propensity score matching. Age, sex, surgical risk, operation year, and proportion of emergency operations were matched (Figure 1).

**Fig 1.**
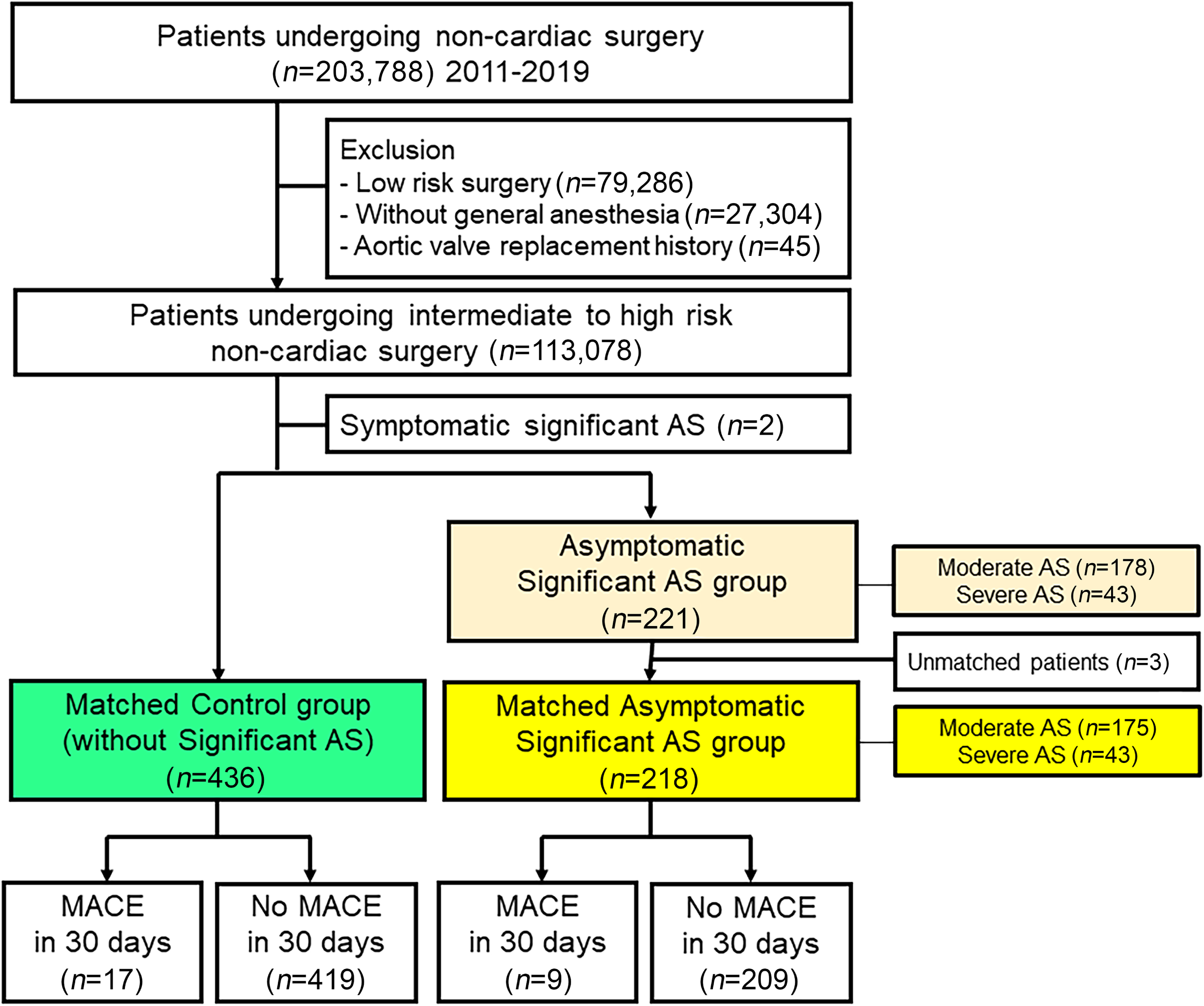
Study flow. Abbreviations: AS, aortic stenosis; MACE, major cardiovascular event; PACE, perioperative cardiovascular events

### Definition and Study Endpoint

All preoperative data, such as demographic information, comorbidity, and laboratory tests, were assessed by independent researchers who were blinded to perioperative events. Perioperative events were assessed by reviewing in-hospital progress notes, nursing charts, discharge notes, and results from cardiac examinations.

The primary outcome was major adverse cardiovascular events (MACE), a composite outcome of heart failure, stroke, myocardial infarction (MI), arrest, and cardiac death within 30 days after surgery. The secondary outcome was perioperative adverse cardiovascular events (PACE), a composite of pulmonary embolism, arrhythmia, heart failure, stroke, MI, arrest, and cardiac death within 30 days of surgery. Following the Fourth Universal Definition of Myocardial Infarction, MI was defined as cardiac marker elevation with the presence of symptoms or new electrocardiographic alterations compatible with MI.^17^ Heart failure was defined when the patient exhibited new or worsening symptoms on presentation, received treatment initiation or intensification specifically for heart failure, or showed objective evidence of new or worsening heart failure.^18^ Arrhythmia included rapid atrial fibrillation, ventricular tachycardia, and bradycardia that necessitated treatment with an antiarrhythmic medication, an electrical shock, or temporary cardiac pacing. Stroke is characterized by a loss of neurological function brought on by an ischemia or hemorrhagic event and manifested by symptoms that last for at least 24 hours.

A representative analysis is as follows. First, to investigate the effect of significant AS on adverse events after surgery, MACE and PACE occurrences were compared in the control group and asymptomatic significant AS group. Second, to determine the factors affecting the occurrence of MACE and PACE in patients with asymptomatic significant AS, patients with and without MACE and PACE were compared and multivariable logistic analysis was performed in patients with asymptomatic significant AS. The study protocol was approved by the Institutional Review Board of SMC and the requirement for informed consent from patients was waived. This study was conducted according to the principles of the Declaration of Helsinki (IRB No. 2021-06-078).

### Echocardiography

Two-dimensional echocardiography was performed under hemodynamically stable conditions using commercially available equipment and analyzed according to the current guidelines.^19^ The evaluation of AS was based on the recommendations of the Echocardiographic Assessment of AS.^15^ Flow acceleration in the aortic valve (AV) was evaluated using the apical 5-chamber view and right parasternal view with continuous wave Doppler; the maximum velocity and pressure gradient values were used for analysis. The velocity time integral of the left ventricular outflow track was evaluated on the apical 5-chamber view with pulsed wave Doppler. The left ventricular outflow track diameter was defined as the mid-systolic inner edge-to-inner edge diameter within 1 cm of the aortic orifice at the parasternal long-axis view. For two-dimensional strain analysis, images were analyzed offline using vendor-independent 2D Cardiac Performance Analysis software, version 1.1.3 (Tom Tec Imaging Systems, Munich, Germany). The endocardial border of the left ventricle (LV) was manually traced from three apical views (apical 2-, 3-, and 4-chamber views) and averaged to obtain the LV global longitudinal strain (GLS).^20^ The endocardial border of the left atrium (LA) was manually traced, similarly to the LV. Starting at the endocardial border of the mitral annulus, the LA endocardial borders were traced using the four and two-chamber views, extrapolating the pulmonary veins and LA appendage orifices, up to the opposing mitral annulus side.^21^ In this study, the LA reservoir strain was used in the analysis. The cardiac damage stage was evaluated by adding the LA and LV strain value based on the established model.^8,9,22^ The cardiac damage is stage 4 if RV damage is present, stage 3 if pulmonary vasculature or tricuspid damage is visible, stage 2 if LA or mitral damage is visible, stage 1 if LV damage is visible, and stage 0 if there is no sign of cardiac damage.

### Statistical Analysis

Binary logistic regression was used for propensity score estimation with a caliper. The caliper size was set to 0.2 of the standard deviation and the number of control groups was set to double the significant AS group. The covariate balance after the propensity score matching was assessed by calculating the absolute standardized mean differences. Standardized mean differences were within 10% across all matched covariates, suggesting successful balance achievement between significant AS and control groups. Continuous variables are presented as mean ± standard deviation or median (interquartile range, IQR) using Student’s t-test or Mann-Whitney U test. The Shapiro-Wilk test was used for the normality assumption of continuous variables. Categorical variables were compared between groups using the chi-square test or Fisher’s exact test and presented as numbers and relative frequencies (%). A stepwise multivariable Cox analysis approach was used to eliminate insignificant factors among the factors that were considered clinically important or showed differences at baseline. The optimal cutoff values of the echocardiographic parameters for predicting post-operative outcomes were calculated to maximize the product of sensitivity and specificity using receiver operating characteristic (ROC) curves.

All *P*-values were two-sided and *P*-values < 0.05 were considered statistically significant. Statistical analyses were performed using R Statistical Software (version 4.1.0; R Foundation for Statistical Computing, Vienna, Austria).

## RESULTS

In this study, the median age of 221 patients with asymptomatic significant AS was 74.0 [69.0-79.0] years. Emergent surgery occurred in 15/221 (6.8%) and high-risk surgery in 27/221 (12.2%). In the significant AS group, the median AV V max was 3.3 [3.0-3.6] m/s, mean PG was 23.3 [19.6-30.5] mmHg, and aortic valve area (AVA) was 1.2 [1.0-1.4] cm^2^. The mean age of patients with severe AS was 74.9 ± 6.5 years, AV V max was 4.2 ± 0.4 m/s, mean PG was 39.7 ± 8.6 mmHg, and AVA was 0.9 ± 0.2 cm^2^. Among the 221 patients, 17.6% (39/221) were classified as cardiac damage stage 0, 16.7% (37/221) as stage 1, 63.8% (141/221) as stage 2, 1.8% (4/221) as stage 3, and no patients were categorized as stage 4.

### Baseline Characteristics Comparison Between Matched Control Group and Matched Asymptomatic Significant AS Group

When comparing the matched control group with significant AS, there was no significant difference except for the frequency of hypertension and diabetes and the level of anemia (Table S1). When comparing the echocardiography parameters between the matched control group and the matched asymptomatic significant AS group, left ventricular end diastolic dimension, left ventricular mass index, E/e’, left atrium volume index, and right ventricular systolic pressure values were significantly higher in the AS group and mitral annulus tissue velocity was significantly lower in the AS group. There was no significant difference in the left ventricular ejection fraction (LVEF) between the two groups (Table S2).

### Clinical Outcome Comparison Between Matched Control Group and Matched Asymptomatic Significant AS Group

The perioperative outcome was divided into three groups of matched populations: control, moderate, and severe AS. In the control group, MACE events were 3.9% (17/436), 4% (7/175) in the moderate AS group, and 4.7% (2/43) in the severe AS group. The PACE event was 13.5% (59/436) in the control group, 16.6% (29/175) in the moderate AS group, and 23.3% (10/43) in the severe AS group (Table 1). There was no significant difference between the three groups in the ratio of primary and secondary outcomes, MACE (*P*=0.971) and PACE (*P*=0.185). Hard outcomes such as cardiac death, arrest, MI, and stroke were also not significantly different. The only event with a significant difference between the three groups was post-operative arrhythmia events (*P=*0.038) (Figure 2).

**Fig 2.**
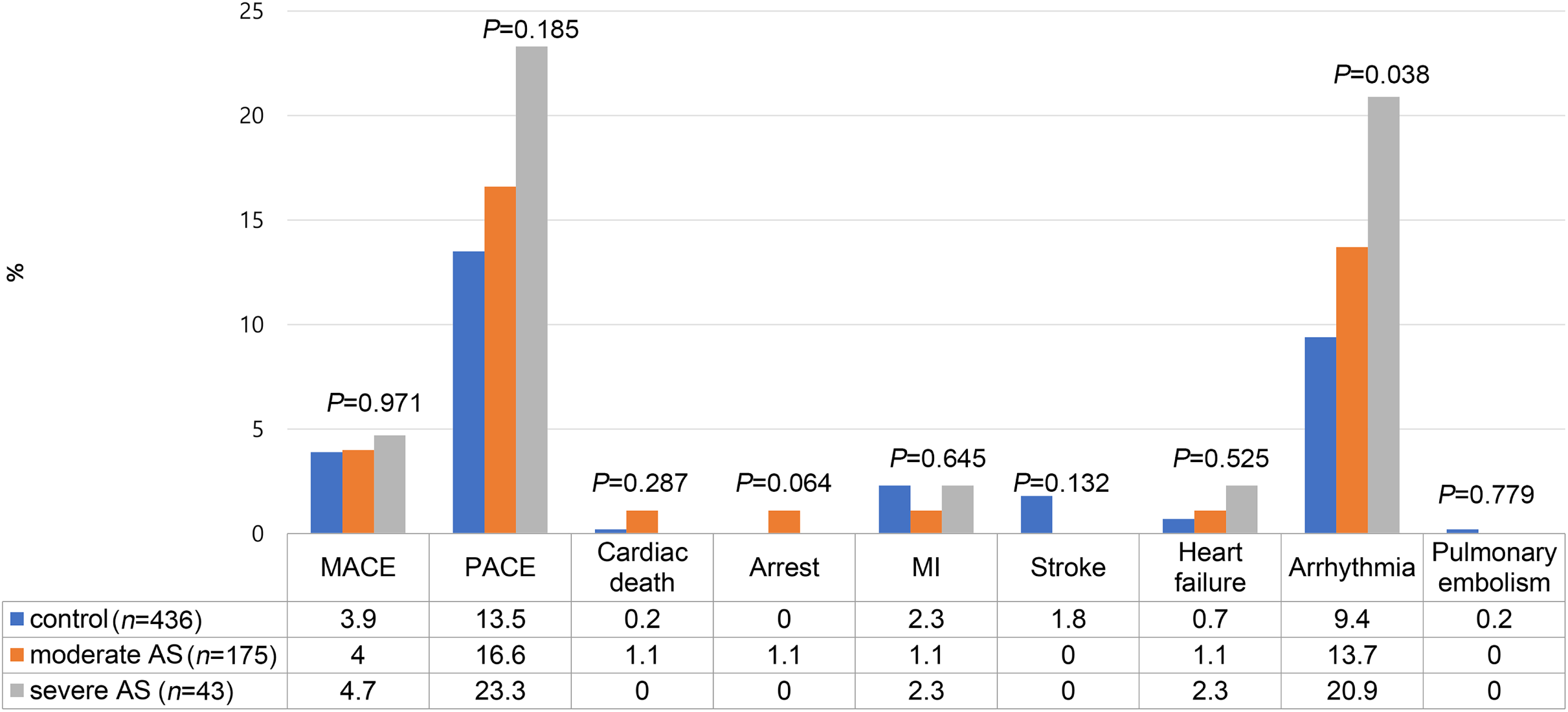
Perioperative outcomes in 30 days. Abbreviations: AS, aortic stenosis; MACE, major cardiovascular event; PACE, perioperative cardiovascular events

**Table 1.** Post-Operative Outcome Comparison Between Significant AS Patient Group and Matched Control Group in 30 Days.

| Variable | Control group<br>(n=436) | Significant AS group |  |  | P-value<br>(Moderate AS<br>vs Control) | P-value<br>(Severe AS<br>vs Control) | P-value<br>(Significant AS<br>vs Control) |
| --- | --- | --- | --- | --- | --- | --- | --- |
|  |  | Moderate AS<br>(n=175) | Severe AS<br>(n=43) | Total<br>(n =218) |  |  |  |
| Composite clinical outcome within 30 days |  |  |  |  |  |  |  |
| MACE | 17 (3.9%) | 7 (4.0%) | 2 (4.7%) | 9 (4.1%) | 1.000 | 1.000 | 1.000 |
| PACE | 59 (13.5%) | 29 (16.6%) | 10 (23.3%) | 39 (17.9%) | 0.401 | 0.132 | 0.175 |
| Clinical outcome within 30 days |  |  |  |  |  |  |  |
| Cardiac death | 1 (0.2%) | 2 (1.1%) | 0 (0.0%) | 2 (0.9%) | 0.412 | 1.000 | 0.539 |
| Arrest | 0 (0.0%) | 2 (1.1%) | 0 (0.0%) | 2 (0.9%) | 0.146 | NA | 0.211 |
| MI | 10 (2.3%) | 2 (1.1%) | 1 (2.3%) | 3 (1.4%) | 0.546 | 1.000 | 0.620 |
| Stroke | 8 (1.8%) | 0 (0.0%) | 0 (0.0%) | 0 (0.0%) | 0.158 | 0.786 | 0.102 |
| Heart failure | 3 (0.7%) | 2 (1.1%) | 1 (2.3%) | 3 (1.4%) | 0.946 | 0.805 | 0.664 |
| Arrhythmia | 41 (9.4%) | 24 (13.7%) | 9 (20.9%) | 33 (15.1%) | 0.156 | **0.036** | **0.040** |
| Pulmonary embolism | 1 (0.2%) | 0 (0.0%) | 0 (0.0%) | 0 (0.0%) | 1.000 | 1.000 | 1.000 |

Data are presented as n (%). The values in bold indicate statistical significance ( $P<0.05$ ).
AS, aortic stenosis; MACE, late diastolic filling velocity; MI, myocardial infarction; PACE, Perioperative adverse cardiovascular events.

When the three groups were analyzed separately, namely moderate AS versus control, severe AS versus control, and significant AS versus control, no significant differences were observed in MACE and PACE events. The only significant difference observed was a higher occurrence of arrhythmia events in severe AS compared to the control group (20.9% at severe AS, 9.4% at control group, *P*=0.036) and in significant AS compared to the control group (15.1% at severe AS, 9.4% at control group, *P*=0.040) (Table 1).

### Factors Associated With Clinical Outcomes in Patient With Asymptomatic Significant AS

When the factors related to MACE and PACE were analyzed by logistic regression in the total asymptomatic significant AS population (n=221). Significant factors with MACE in the univariable logistic regression were the presence of underlying CAD (OR 5.389, 95% CI [1.242-23.382], *P*=0.025), hemoglobin level (OR 0.640, 95% CI [0.428-0.959], *P*=0.031), and cardiac damage stage (OR 8.611, 95% CI [1.426-51.993], *P=*0.019). When multivariable logistic regression was performed with CAD, hemoglobin level, and cardiac damage stage, significant factors with MACE were CAD (OR 6.256, 95% CI [1.258-31.115], *P*=0.026) and cardiac damage stage (OR 9.241, 95% CI [1.314-64.976], *P*=0.025) (Figure 3A, Table S3).

**Fig 3.**
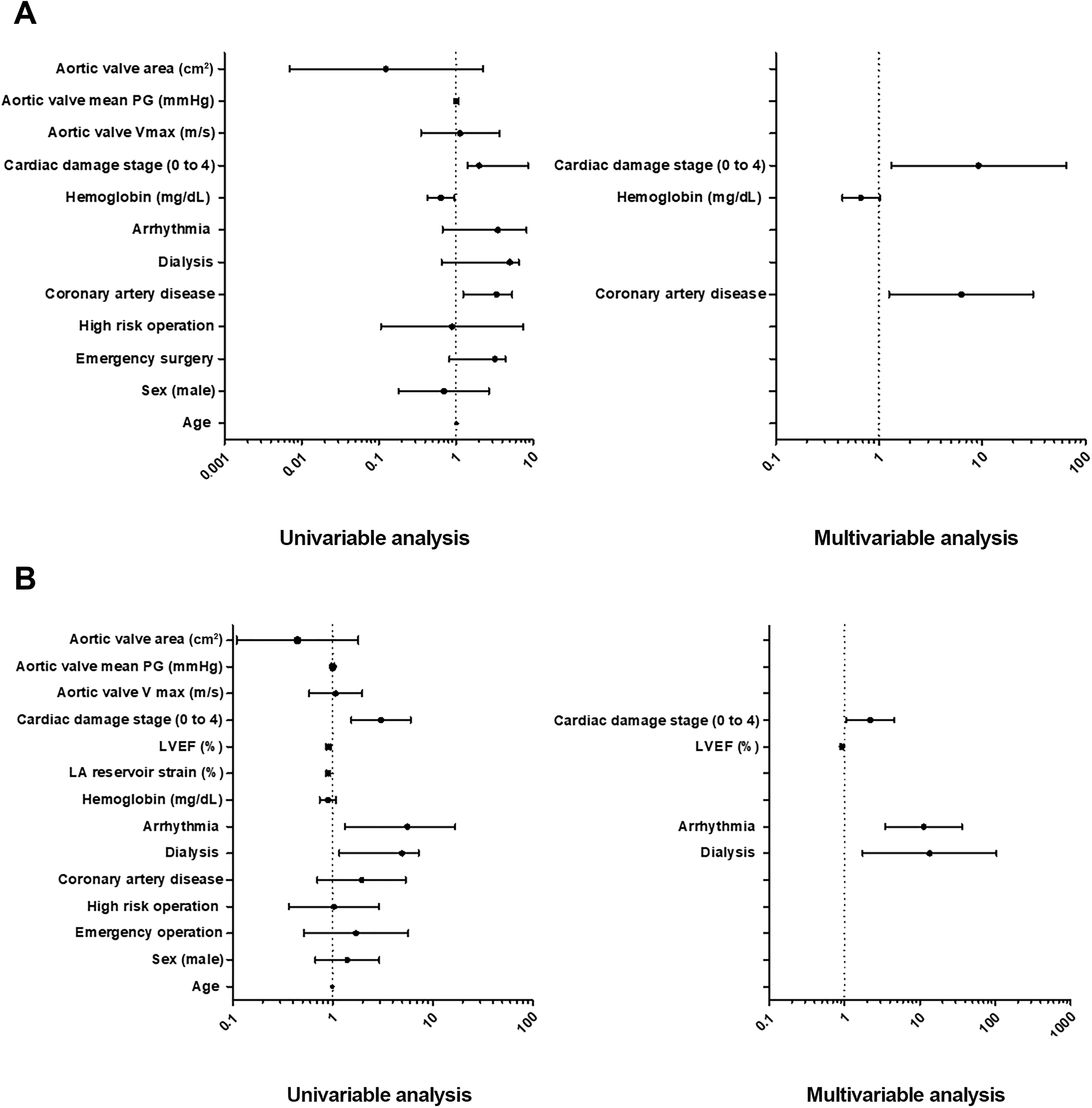
(A) Univariable and multivariable risk factor analysis for MACE events. (B) Univariable and multivariable risk factor Analysis for PACE events. Abbreviations: CI, confidence interval; OR, odds ratio; PG, pressure gradient; MACE, major cardiovascular event; PACE, perioperative cardiovascular events

Factors significantly associated with PACE in univariable logistic regression were dialysis (OR 7.257, 95% CI [1.171-44.955], *P*=0.033), arrhythmia (OR 16.948, 95% CI [5.596-51.329], *P*=<0.001), LA reservoir strain (OR 0.908, 95% CI [0.864-0.953], *P*=<0.001), LVEF (OR 0.908, 95% CI [0.863-0.955], *P*=<0.001), and cardiac damage stage (OR 3.046, 95% CI [1.543-6.009], *P*=0.001). When multivariable analysis factors were selected through backward elimination, dialysis, arrhythmia, LVEF, and cardiac damage stage remained, and all four factors were significantly associated with PACE. The OR of dialysis was 13.486 (95% CI [1.742-104.371], *P*=0.013), the OR of arrhythmia was 11.219 (95% CI [3.474-36.231], *P*<0.001), the OR of LVEF was 0.928 (95% CI [0.882-0.977], *P=*0.004), and the OR of cardiac damage stage was 2.199 (95% CI [1.055-4.584], *P*=0.036) (Figure 3B, Table S4).

When the cutoff point for increasing MACE and PACE was obtained by using a ROC curve, MACE increased rapidly at an LVEF of less than 62.5%, LV GLS was more than - 13.6%, and LA reservoir strain less than 25.9%. Similarly, PACE increased at an LVEF of less than 61.7%, LV GLS was more than -16.7%, and the LA reservoir strain was less than 24.3%. The AUC values of all three parameters were statistically significant (Figure 4).

**Fig 4.**
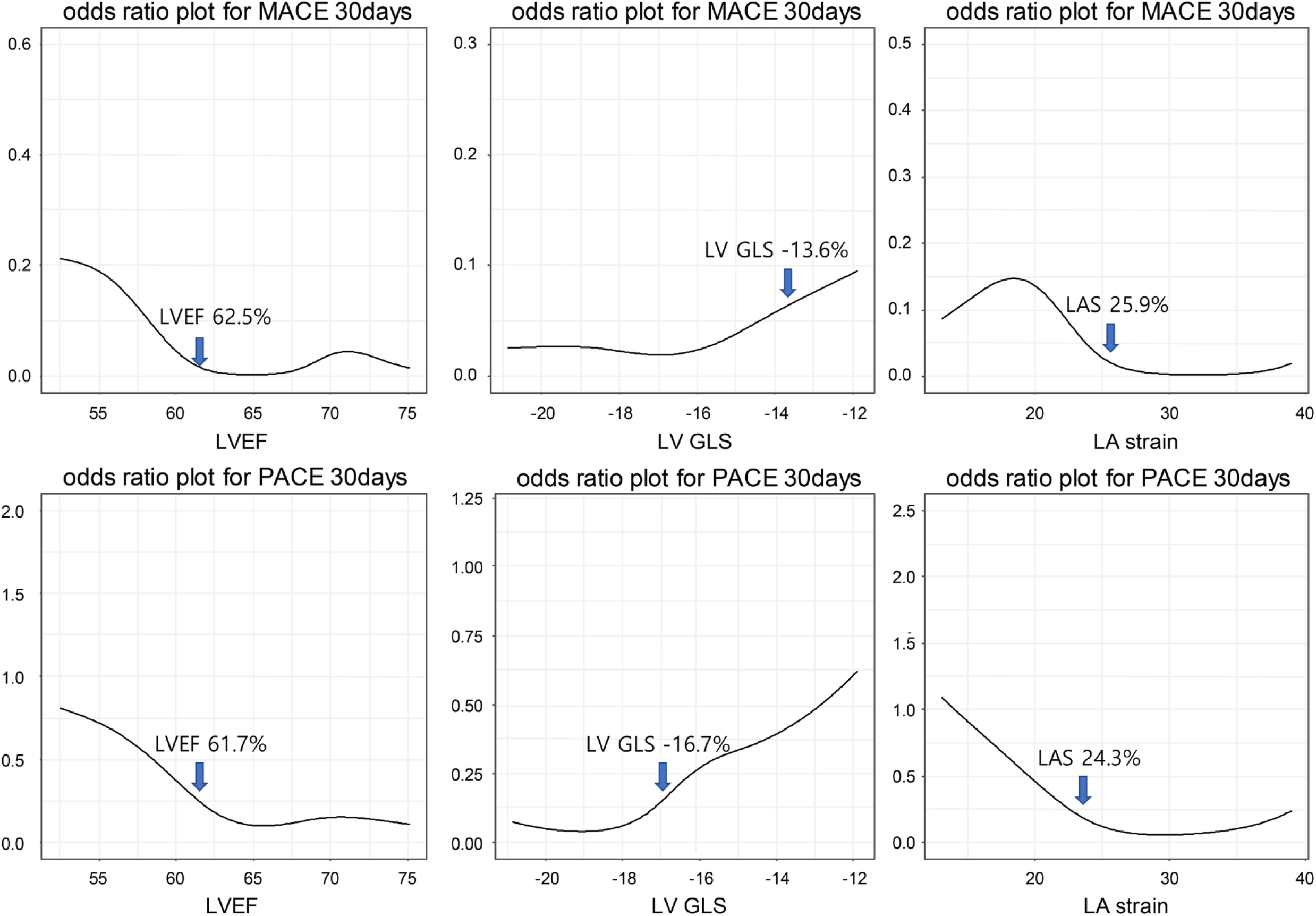
Cutoff value of echo parameters increasing perioperative complication risk. Abbreviations: LAS, left atrium strain; LVEF, left ventricular ejection fraction; LV GLS, left ventricle global longitudinal strain; MACE, major cardiovascular event; PACE, perioperative cardiovascular events.

### Subgroup Analysis in Patients With Severe AS

When subgroup analysis was performed on 43 patients with severe AS, the two patients with MACE had no specific underlying disease other than AS, and their ages were within the 95% confidence interval of the control group (Table 2). In both patients with MACE, the pre-operative N-terminal pro brain natriuretic peptide level was higher than the 95% confidence interval of the control group, and the absolute values of LVEF, LV GLS, and LA strain were lower than the 95% confidence interval of the control group. Both patients had an advanced cardiac damage stage 2 and 3, respectively (Table 3). Both MACE events happened after vascular surgery.

**Table 2.** Baseline Characteristics Comparison Between MACE and Non-MACE Group in Asymptomatic Severe AS Patients.

|  | Without MACE (n=41) | Patient 1 | Patient 2 | Total (n=43) |
| --- | --- | --- | --- | --- |
| **Demographic information** |  |  |  |  |
| Age (years) | 75.0 ± 6.5 | 78 | 68 | 74.9 ± 6.5 |
| Sex (male) | 30 (73.2%) | Male | Female | 31 (72.1%) |
| **Comorbidity** |  |  |  |  |
| Hypertension | 26 (63.4%) | No | No | 26 (60.5%) |
| Diabetes | 9 (22.0%) | No | No | 9 (20.9%) |
| Chronic kidney disease | 2 (4.9%) | No | No | 2 (4.7%) |
| Stroke history | 3 (7.3%) | No | No | 3 (7.0%) |
| Coronary artery disease | 4 (9.8%) | No | No | 4 (9.3%) |
| Heart failure | 1 (2.4%) | No | No | 1 (2.3%) |
| Atrial fibrillation | 3 (7.3%) | No | No | 3 (7.0%) |
| PAOD | 1 (2.4%) | No | No | 1 (2.3%) |
| **Operative information** |  |  |  |  |
| Emergency surgery | 5 (12.2%) | No | Yes | 6 (14.0%) |
| High-risk surgery | 8 (19.5%) | No | No | 8 (18.6%) |
| Operation time (minutes) | 153.6 ± 72.6 | 123 | **72** | 151.0 ± 72.0 |

Data are presented as mean ± standard deviation, median [25–75 percentiles], or n (%). The values in bold indicate value outside the 95% confidence interval.
MACE, major adverse cardiovascular events; PAOD, peripheral artery occlusive disease.

**Table 3.**
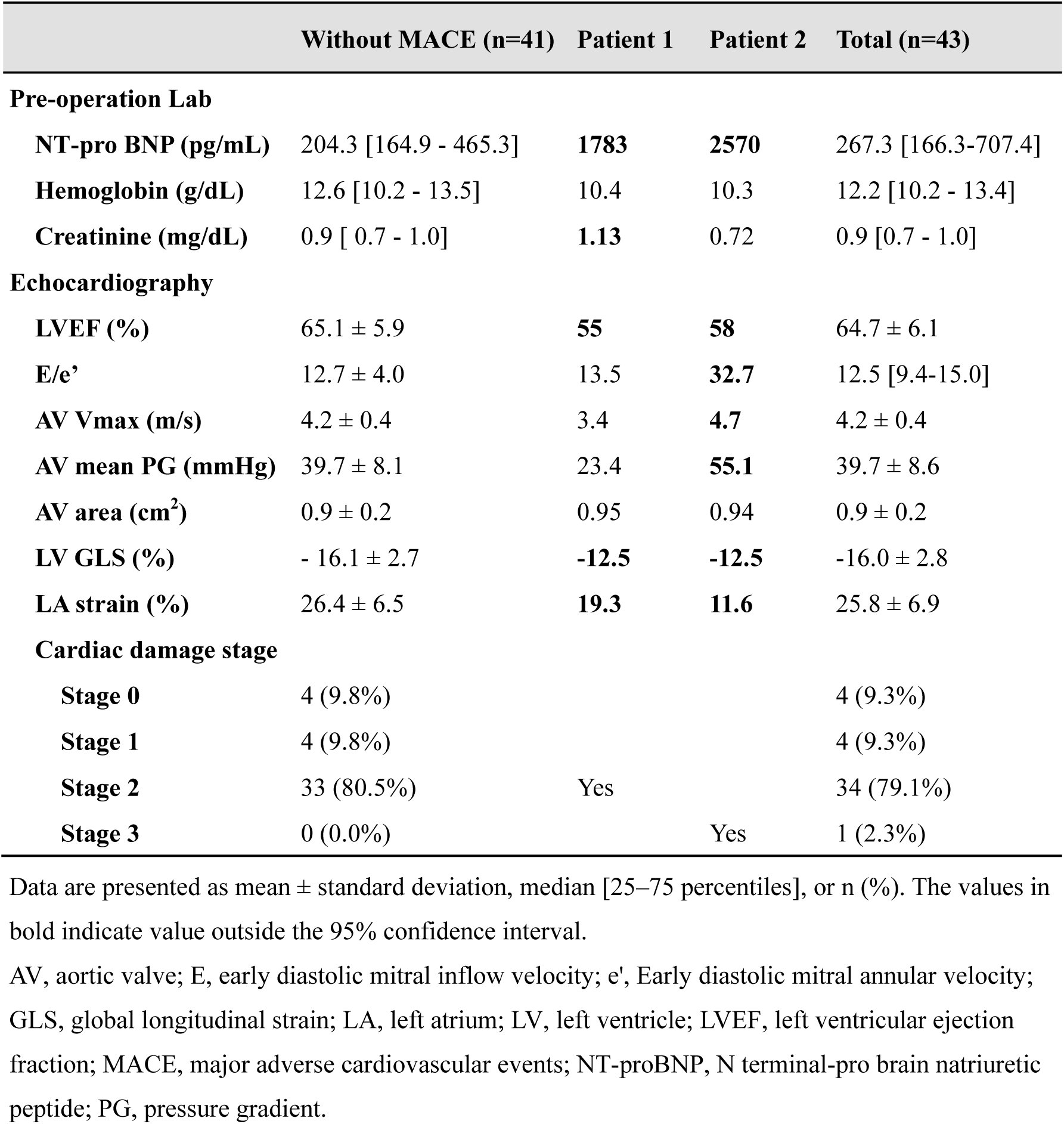
Baseline Laboratory and Echocardiographic Parameters Comparison Between MACE and Non-MACE Group in Asymptomatic Severe AS Patients.

## DISCUSSION

This study demonstrated that patients with asymptomatic significant AS did not experience significantly higher perioperative complications during non-cardiac surgery when compared to the control group without AS. Notably, our findings suggest that the extent of cardiac damage resulting from AS had a greater influence on perioperative complications than the severity of the AS itself. While our study underscores the importance of individualized risk stratification and careful preoperative evaluation in patients with severe asymptomatic AS who require non-cardiac surgery, it is important to note that patients who need surgery should not avoid it solely based on the presence of AS. Rather, the decision to undergo surgery should be made after a comprehensive evaluation of overall health status, comorbidities, and surgical risk, with input from a multidisciplinary team comprising cardiologists, anesthesiologists, and surgeons. Such an approach can help identify patients who may benefit from surgery while minimizing the potential risks.

### Decreased Perioperative Complications Over Time

Comparing this study with that of Tashiro et al. 10 years ago,^7^ the MACE incidence rate of patients with asymptomatic severe AS has decreased from 12% to 4.7%. Similarly in the control group, the MACE incidence rate, which was 10.5% in the past, decreased to 3.9%.^7^ Furthermore, in the present study, mortality or perioperative myocardial infarction (MI) was observed in 2.3% (1/43) of patients with asymptomatic severe AS, which is notably lower compared to the 4.7% incidence reported in the prior study by Agarwal et al.^5^ In summary, it appears that perioperative complications of non-cardiac surgery are diminishing over time, whether in patients with concomitant AS or those without.

### Comparison of Perioperative Complications Between Patients With Asymptomatic Significant AS and Patients Without AS

Although the asymptomatic severe AS group and the control group were not compared directly in the study by Tashiro et al,^7^ it is noteworthy that the incidence of perioperative events was remarkably higher in the symptomatic AS group than in the other group. And there was little numerical difference between the asymptomatic severe AS group and control group. Likewise in the Agarwal et al. study,^5^ there was no significant difference in perioperative outcomes between the asymptomatic severe AS and control groups, whereas there was a significant difference in outcomes between the symptomatic AS and control groups. Consequently, upon exclusion of symptomatic patients, there was no statistically significant difference in the perioperative complication rate of non-cardiac surgery between patients with AS and those without AS, both a decade ago and at present.

### Predisposing Factors of Perioperative Complication in Patients With AS

In this study, we found that CAD and cardiac damage stage were independently associated with MACE, while dialysis, underlying arrhythmia, LVEF, and cardiac damage stage were independently associated with PACE. Although anemia was a prognostic factor for MACE in univariable analysis, its significance disappeared in multivariable analysis, suggesting that anemia reflects comorbidity, rather than being an independent prognostic factor in this study. CAD is often associated with AS and has been identified as a risk factor for perioperative mortality or MI in previous studies.^5^ Even without CAD, AS can impair coronary vasodilatation and cause a mismatch between oxygen supply and demand, leading to myocardial ischemia.^23^ CAD can accelerate this myocardial ischemia. In this study, dialysis was the main risk factor for PACE, probably due to combined cardiovascular risk factors and diastolic dysfunction in dialysis patients.^24–26^ For reference, dialysis patients have a high perioperative mortality rate in general elective surgery, even without AS.^26^

### Importance of Cardiac Remodeling in Perioperative Complication in Patients With AS

In this study, advanced cardiac damage stage was identified as an independent and important risk factor for MACE and PACE. Patients with AS who experience congestive heart failure, atrial fibrillation, and elevated right ventricular systolic pressure are at higher risk of post-operative cardiovascular events.^5,7,10,24,27^ These factors are all components of the cardiac damage stage in patients with AS. Hemodynamic changes during surgery, such as tachycardia, anemia, stress, and sympathomimetic drug effects, can lead to myocardial ischemia and increased ventricular filling pressure.^28,29^ Patients with advanced cardiac damage stage may be more vulnerable to these changes due to a smaller reservoir. For this reason, heart failure has been considered a representative risk factor for perioperative complications, as stated in the 2014 ACC/AHA guideline^14^ and the American College of Surgeons National Surgical Quality Improvement Program model.^30^ This study is the first to establish a significant relationship between the recently proposed cardiac damage stage and perioperative outcomes in patients with AS. In this study, each stage of cardiac damage was associated with a 9.3-fold increase in the risk of MACE and a 2.2-fold increase in the risk of PACE. Additional studies with a larger sample size may be necessary to obtain a more reliable odds ratio.

Several studies have reported that parameters such as mean pressure gradient, aortic valve area (AVA), and AV maximum velocity (AV Vmax) in patients with AS are associated with perioperative outcomes.^4,7,31^ However, these parameters were only found to be significant in univariable analysis and not in multivariable analysis.^7^ Additionally, it was observed that patients with worse AV hemodynamics had lower LVEF.^4^ In our study, the univariable logistic regression analysis revealed that all three AV parameters (AV Vmax, AV mean PG, and AVA) had no statistically significant relationship with MACE and PACE. On the other hand, in multivariable analysis, LVEF and cardiac damage stage appeared as significant factors. These findings suggest that cardiac damage is more crucial than AV hemodynamics in perioperative outcomes. Agarwal et al. also reported a similar result, where there was no significant difference in AVA or mean pressure gradient, but LVEF and severity of mitral regurgitation were significantly different between patients with and without primary outcomes. Our study aligns with the 2022 ESC guideline, which recommends the consideration of LVEF in the decision-making process for surgery in patients with severe AS.^32^ However, whether the normal LVEF threshold for AS should be 50%, 55%, or 60% remains controversial.^33–35^ Our study suggests that the myocardial status has a more significant effect on perioperative outcomes than the actual degree of AS. Nevertheless, future studies are needed to determine the appropriate LVEF criteria for surgery.

### Limitation

This study, conducted at a single center from 2011 to 2019, has the advantage of confirming the perioperative outcome of patients with asymptomatic significant AS but has several limitations. First, because patients with significant AS were found among those who underwent echocardiography before surgery, there is a possibility that those who did not undergo echocardiography or those who did not diagnosed before surgery were not included. However, according to the strategy of the Department of Anesthesiology at this hospital, echocardiography is performed on all patients aged 65 years or older, and the average age of Korean patients diagnosed with AS is 69 years old,^2^ therefore, most of the patients with degenerative AS are expected to be included. Second, there may be limitations in generalization because the analysis is for tertiary hospital surgery where experienced medical staff deal with patients with advanced diseases. Third, there are limitations in retrospective studies. Unreported values may exist among the variables, and there is a possibility that the outcome may be missed in the case of patients who are discharged within 30 days and have no follow-up. However, by double-checking the events through meticulous chart review, we tried to ensure that there were no missing variables or outcomes. Fourth, it is possible that symptoms caused by AS were under-reported in patients with poor functional status. In our hospital, when non-cardiac surgery is performed on a patient with significant AS, a valve specialist must conduct a preoperative consultation. Therefore, it is assumed that there would have been no difficulty in determining whether the symptoms were caused by AS during the process of history taking.

## CONCLUSIONS

There was no significant difference in major post-operative cardiovascular events between patients with asymptomatic significant AS and the control group. However, we found that the advanced cardiac damage stage in significant AS is an important factor in perioperative risk assessment for non-cardiac surgery. These findings suggest the need for careful pre-operative evaluation and individualized risk stratification in patients with significant AS undergoing non-cardiac surgery, particularly those with more advanced disease.

## Data Availability

The data that support the findings of this study are available from the corresponding author on reasonable request.

## Nonstandard Abbreviations and Acronyms

AS: Aortic stenosis
AV: Aortic valve
CAD: Coronary artery disease
GLS: Global longitudinal strain
LA: Left atrium
LV: Left ventricle
LVEF: Left ventricular ejection fraction
MACE: Major adverse cardiovascular events
MI: Myocardial infarction
PACE: Perioperative adverse cardiovascular events
PG: Pressure gradient

## REFERENCES

1. Rezzoug N, Vaes B, de Meester C, Degryse J, Van Pottelbergh G, Mathei C, Adriaensen W, Pasquet A, Vanoverschelde JL. The clinical impact of valvular heart disease in a population-based cohort of subjects aged 80 and older. BMC Cardiovasc Disord. 2016;16:7. doi: 10.1186/s12872-016-0184-8

2. Jang SY, Park SJ, Kim EK, Park SW. Temporal trends in incidence, prevalence, and death of aortic stenosis in Korea: a nationwide population-based study. ESC Heart Fail. 2022;9:2851–2861. doi: 10.1002/ehf2.13957

3. Banovic MD, Vujisic-Tesic BD, Kujacic VG, Callahan MJ, Nedeljkovic IP, Trifunovic DD, Aleksandric SB, Petrovic MZ, Obradovic SD, Ostojic MC. Coronary flow reserve in patients with aortic stenosis and nonobstructed coronary arteries. Acta Cardiol. 2011;66:743–749. doi: 10.1080/ac.66.6.2136958

4. Kertai MD, Bountioukos M, Boersma E, Bax JJ, Thomson IR, Sozzi F, Klein J, Roelandt JR, Poldermans D. Aortic stenosis: an underestimated risk factor for perioperative complications in patients undergoing noncardiac surgery. Am J Med. 2004;116:8–13. doi: 10.1016/j.amjmed.2003.07.012

5. Agarwal S, Rajamanickam A, Bajaj NS, Griffin BP, Catacutan T, Svensson LG, Anabtawi AG, Tuzcu EM, Kapadia SR. Impact of aortic stenosis on postoperative outcomes after noncardiac surgeries. Circ Cardiovasc Qual Outcomes. 2013;6:193–200. doi: 10.1161/CIRCOUTCOMES.111.000091

6. Li G, Warner M, Lang BH, Huang L, Sun LS. Epidemiology of anesthesia-related mortality in the United States, 1999-2005. Anesthesiology. 2009;110:759–765. doi: 10.1097/aln.0b013e31819b5bdc

7. Tashiro T, Pislaru SV, Blustin JM, Nkomo VT, Abel MD, Scott CG, Pellikka PA. Perioperative risk of major non-cardiac surgery in patients with severe aortic stenosis: a reappraisal in contemporary practice. Eur Heart J. 2014;35:2372–2381. doi: 10.1093/eurheartj/ehu044

8. Genereux P, Pibarot P, Redfors B, Mack MJ, Makkar RR, Jaber WA, Svensson LG, Kapadia S, Tuzcu EM, Thourani VH, et al. Staging classification of aortic stenosis based on the extent of cardiac damage. Eur Heart J. 2017;38:3351–3358. doi: 10.1093/eurheartj/ehx381

9. Tastet L, Tribouilloy C, Marechaux S, Vollema EM, Delgado V, Salaun E, Shen M, Capoulade R, Clavel MA, Arsenault M, et al. Staging Cardiac Damage in Patients With Asymptomatic Aortic Valve Stenosis. J Am Coll Cardiol. 2019;74:550–563. doi: 10.1016/j.jacc.2019.04.065

10. Raymer K, Yang H. Patients with aortic stenosis: cardiac complications in non-cardiac surgery. Can J Anaesth. 1998;45:855–859. doi: 10.1007/BF03012219

11. Torsher LC, Shub C, Rettke SR, Brown DL. Risk of patients with severe aortic stenosis undergoing noncardiac surgery. Am J Cardiol. 1998;81:448–452. doi: 10.1016/s0002-9149(97)00926-0

12. Calleja AM, Dommaraju S, Gaddam R, Cha S, Khandheria BK, Chaliki HP. Cardiac risk in patients aged >75 years with asymptomatic, severe aortic stenosis undergoing noncardiac surgery. Am J Cardiol. 2010;105:1159–1163. doi: 10.1016/j.amjcard.2009.12.019

13. Kristensen SD, Knuuti J. New ESC/ESA Guidelines on non-cardiac surgery: cardiovascular assessment and management. Eur Heart J. 2014;35:2344–2345. doi: 10.1093/eurheartj/ehu285

14. Fleisher LA, Fleischmann KE, Auerbach AD, Barnason SA, Beckman JA, Bozkurt B, Davila-Roman VG, Gerhard-Herman MD, Holly TA, Kane GC, et al. 2014 ACC/AHA guideline on perioperative cardiovascular evaluation and management of patients undergoing noncardiac surgery: a report of the American College of Cardiology/American Heart Association Task Force on practice guidelines. J Am Coll Cardiol. 2014;64:e77–137. doi: 10.1016/j.jacc.2014.07.944

15. Baumgartner H, Hung J, Bermejo J, Chambers JB, Edvardsen T, Goldstein S, Lancellotti P, LeFevre M, Miller F, Jr., Otto CM. Recommendations on the Echocardiographic Assessment of Aortic Valve Stenosis: A Focused Update from the European Association of Cardiovascular Imaging and the American Society of Echocardiography. J Am Soc Echocardiogr. 2017;30:372–392. doi: 10.1016/j.echo.2017.02.009

16. Strange G, Stewart S, Celermajer D, Prior D, Scalia GM, Marwick T, Ilton M, Joseph M, Codde J, Playford D, et al. Poor Long-Term Survival in Patients With Moderate Aortic Stenosis. J Am Coll Cardiol. 2019;74:1851–1863. doi: 10.1016/j.jacc.2019.08.004

17. Thygesen K, Alpert JS, Jaffe AS, Chaitman BR, Bax JJ, Morrow DA, White HD, Executive Group on behalf of the Joint European Society of Cardiology /American College of Cardiology /American Heart Association /World Heart Federation Task Force for the Universal Definition of Myocardial I. Fourth Universal Definition of Myocardial Infarction (2018). J Am Coll Cardiol. 2018;72:2231–2264. doi: 10.1016/j.jacc.2018.08.1038

18. Hicks KA, Tcheng JE, Bozkurt B, Chaitman BR, Cutlip DE, Farb A, Fonarow GC, Jacobs JP, Jaff MR, Lichtman JH, et al. 2014 ACC/AHA Key Data Elements and Definitions for Cardiovascular Endpoint Events in Clinical Trials: A Report of the American College of Cardiology/American Heart Association Task Force on Clinical Data Standards (Writing Committee to Develop Cardiovascular Endpoints Data Standards). J Nucl Cardiol. 2015;22:1041–1144. doi: 10.1007/s12350-015-0209-1

19. Lang RM, Badano LP, Mor-Avi V, Afilalo J, Armstrong A, Ernande L, Flachskampf FA, Foster E, Goldstein SA, Kuznetsova T, et al. Recommendations for cardiac chamber quantification by echocardiography in adults: an update from the American Society of Echocardiography and the European Association of Cardiovascular Imaging. Eur Heart J Cardiovasc Imaging. 2015;16:233–270. doi: 10.1093/ehjci/jev014

20. Voigt JU, Pedrizzetti G, Lysyansky P, Marwick TH, Houle H, Baumann R, Pedri S, Ito Y, Abe Y, Metz S, et al. Definitions for a common standard for 2D speckle tracking echocardiography: consensus document of the EACVI/ASE/Industry Task Force to standardize deformation imaging. J Am Soc Echocardiogr. 2015;28:183–193. doi: 10.1016/j.echo.2014.11.003

21. Badano LP, Kolias TJ, Muraru D, Abraham TP, Aurigemma G, Edvardsen T, D’Hooge J, Donal E, Fraser AG, Marwick T, et al. Standardization of left atrial, right ventricular, and right atrial deformation imaging using two-dimensional speckle tracking echocardiography: a consensus document of the EACVI/ASE/Industry Task Force to standardize deformation imaging. Eur Heart J Cardiovasc Imaging. 2018;19:591–600. doi: 10.1093/ehjci/jey042

22. Sun BJ, Park JH, Lee M, Choi JO, Lee JH, Shin MS, Kim MJ, Jung HO, Park JR, Sohn IS, et al. Normal Reference Values for Left Atrial Strain and Its Determinants from a Large Korean Multicenter Registry. J Cardiovasc Imaging. 2020;28:186–198. doi: 10.4250/jcvi.2020.0043

23. Smucker ML, Tedesco CL, Manning SB, Owen RM, Feldman MD. Demonstration of an imbalance between coronary perfusion and excessive load as a mechanism of ischemia during stress in patients with aortic stenosis. Circulation. 1988;78:573–582. doi: 10.1161/01.cir.78.3.573

24. Fayad A, Ansari MT, Yang H, Ruddy T, Wells GA. Perioperative Diastolic Dysfunction in Patients Undergoing Noncardiac Surgery Is an Independent Risk Factor for Cardiovascular Events: A Systematic Review and Meta-analysis. Anesthesiology. 2016;125:72–91. doi: 10.1097/ALN.0000000000001132

25. Pecoits-Filho R, Bucharles S, Barberato SH. Diastolic heart failure in dialysis patients: mechanisms, diagnostic approach, and treatment. Semin Dial. 2012;25:35–41. doi: 10.1111/j.1525-139X.2011.01011.x

26. Palamuthusingam D, Nadarajah A, Pascoe EM, Craig J, Johnson DW, Hawley CM, Fahim M. Postoperative mortality in patients on chronic dialysis following elective surgery: A systematic review and meta-analysis. PLoS One. 2020;15:e0234402. doi: 10.1371/journal.pone.0234402

27. Rohde LE, Polanczyk CA, Goldman L, Cook EF, Lee RT, Lee TH. Usefulness of transthoracic echocardiography as a tool for risk stratification of patients undergoing major noncardiac surgery. Am J Cardiol. 2001;87:505–509. doi: 10.1016/s0002-9149(00)01421-1

28. Haggmark S, Hohner P, Ostman M, Friedman A, Diamond G, Lowenstein E, Reiz S. Comparison of hemodynamic, electrocardiographic, mechanical, and metabolic indicators of intraoperative myocardial ischemia in vascular surgical patients with coronary artery disease. Anesthesiology. 1989;70:19–25. doi: 10.1097/00000542-198901000-00006

29. London MJ, Tubau JF, Wong MG, Layug E, Hollenberg M, Krupski WC, Rapp JH, Browner WS, Mangano DT. The “natural history” of segmental wall motion abnormalities in patients undergoing noncardiac surgery. S.P.I. Research Group. Anesthesiology. 1990;73:644–655. doi: 10.1097/00000542-199010000-00010

30. Cohen ME, Ko CY, Bilimoria KY, Zhou L, Huffman K, Wang X, Liu Y, Kraemer K, Meng X, Merkow R, et al. Optimizing ACS NSQIP modeling for evaluation of surgical quality and risk: patient risk adjustment, procedure mix adjustment, shrinkage adjustment, and surgical focus. J Am Coll Surg. 2013;217:336–346 e331. doi: 10.1016/j.jamcollsurg.2013.02.027

31. Detsky AS, Abrams HB, McLaughlin JR, Drucker DJ, Sasson Z, Johnston N, Scott JG, Forbath N, Hilliard JR. Predicting cardiac complications in patients undergoing non-cardiac surgery. J Gen Intern Med. 1986;1:211–219. doi: 10.1007/BF02596184

32. Halvorsen S, Mehilli J, Cassese S, Hall TS, Abdelhamid M, Barbato E, De Hert S, de Laval I, Geisler T, Hinterbuchner L, et al. 2022 ESC Guidelines on cardiovascular assessment and management of patients undergoing non-cardiac surgery. Eur Heart J. 2022;43:3826–3924. doi: 10.1093/eurheartj/ehac270

33. Lancellotti P, Magne J, Dulgheru R, Clavel MA, Donal E, Vannan MA, Chambers J, Rosenhek R, Habib G, Lloyd G, et al. Outcomes of Patients With Asymptomatic Aortic Stenosis Followed Up in Heart Valve Clinics. JAMA Cardiol. 2018;3:1060–1068. doi: 10.1001/jamacardio.2018.3152

34. Ito S, Miranda WR, Nkomo VT, Connolly HM, Pislaru SV, Greason KL, Pellikka PA, Lewis BR, Oh JK. Reduced Left Ventricular Ejection Fraction in Patients With Aortic Stenosis. J Am Coll Cardiol. 2018;71:1313–1321. doi: 10.1016/j.jacc.2018.01.045

35. Bohbot Y, de Meester de Ravenstein C, Chadha G, Rusinaru D, Belkhir K, Trouillet C, Pasquet A, Marechaux S, Vanoverschelde JL, Tribouilloy C. Relationship Between Left Ventricular Ejection Fraction and Mortality in Asymptomatic and Minimally Symptomatic Patients With Severe Aortic Stenosis. JACC Cardiovasc Imaging. 2019;12:38–48. doi: 10.1016/j.jcmg.2018.07.029

